# Assessment of public attention, risk perception, emotional and behavioural responses to the COVID-19 outbreak: social media surveillance in China

**DOI:** 10.1101/2020.03.14.20035956

**Authors:** Zhiyuan Hou, Fanxing Du, Hao Jiang, Xinyu Zhou, Leesa Lin

## Abstract

**Background:** Using social media surveillance data, this study aimed to assess public attention, risk perception, emotion, and behavioural response to the COVID-19 outbreak in real time.

**Methods:** We collected data from most popular social medias: Sina Weibo, Baidu search engine, and Ali e-commerce marketplace, from 1 Dec 2019 to 15 Feb 2020. Weibo post counts and Baidu searches were used to generate indices assessing public attention. Public intention and actual adoption of recommended protection measures or panic buying triggered by rumours and misinformation were measured by Baidu and Ali indices. Qualitative Weibo posts were analysed by the Linguistic Inquiry and Word Count text analysis programme to assess public emotion responses to epidemiological events, governments’ announcements, and control measures.

**Findings:** We identified two missed windows of opportunity for early epidemic control of the COVID-19 outbreak, one in Dec 2019 and the other between 31 Dec and 19 Jan, when public attention was very low despite the emerging outbreak. Delayed release of information ignited negative public emotions. The public responded quickly to government announcements and adopted recommended behaviours according to issued guidelines. We found rumours and misinformation regarding remedies and cures led to panic buying during the outbreak, and timely clarification of rumours effectively reduced irrational behaviour.

**Interpretation:** Social media surveillance can enable timely assessments of public reaction to risk communication and epidemic control measures, and the immediate clarification of rumours. This should be fully incorporated into epidemic preparedness and response systems.

**Funding:** National Natural Science Foundation of China.

## Research in context

### Evidence before this study

Preparedness experts and the World Health Organisation have urged the integration of social sciences into outbreak response, identified as one of the key research priorities for the response to COVID-19. As a powerful platform for public information sharing and emotional expression, social media’s role in risk communication during emerging infectious diseases outbreaks was first explored during the 2009/2010 A(H1N1) pandemic and again in response to Ebola epidemics. We searched PubMed and EMBASE for all articles published until 9 March, 2020 with the keywords “novel coronavirus,” “CoV,” “COVID-19,” and “SARS-CoV-2.” Only two commentaries called for social media surveillance and managing pubic panic and rumours with information. No social media studies to date have conducted a comprehensive assessment of public attention and awareness, risk perception, and emotion and behavioural responses as they emerge and evolve in real time during the COVID-19 outbreak.

### Added value of this study

Using social media surveillance data, we tracked and assessed real-time public emotional and behavioural responses to government risk communication messages, rumours and misinformation, and major epidemiological events during the first 11 weeks of the COVID-19 outbreak in China. We identified two missed windows of opportunity for early epidemic control in the early stages; we found the delayed release of information ignited negative public sentiment, and that early implementation of containment measures could address public anxiety. Official guidelines, epidemiological information, and rumours can all capture public attention and trigger emotional and behavioural reactions, and timely clarification of rumours can effectively reduce irrational behaviour. This study utilizes rigorous methods and procedures in conducting social media surveillance for COVID-19 with Chinese social media platforms, which can inform future research on social media surveillance for epidemic preparedness and response.

### Implications of all the available evidence

During an outbreak, real-time social media surveillance can enable a timely assessment of public reactions to risk communication, epidemic control measures, and the timely clarification of rumours. Social media surveillance has the potential to track both disease activity before it reaches health care facilities and public concerns and risk perceptions as they emerge and evolve in real time. Future research should explore the epidemiological information shared on these platforms and examine the role of social media in early detection and early warning during epidemic preparedness and response. Findings from this study demonstrate that social media surveillance should be fully incorporated into epidemic preparedness and response system.

## Introduction

In early Dec 2019, novel coronavirus disease 2019 (COVID-19) emerged in Wuhan city, and spread rapidly across China.^1-3^ Since 23 Jan 2020, the Chinese government has put Wuhan and several nearby cities under quarantine and implemented containment measures to slow down community transmission. As of 25 Jan, 30 provinces (autonomous regions, municipalities), excluding Tibet, had launched level 1 major public health emergency responses.^4^ On 31 Jan (Beijing Time), WHO declared the outbreak a public health emergency of international concern (PHEIC). On 2 Mar, a total of 80,302 cases have been reported in China, and the outbreak has spread to sixty-four other countries with 8,993 cases. ^4,5^

During an epidemic, understanding how critical information about the health threat is disseminated and how the public accesses, processes and uses this information is crucial. ^6,7^ The risks and uncertainties of emerging infectious diseases may arouse public emotion and change behaviour in either constructive (e.g. taking up personal hand hygiene and avoiding mass gatherings) or disruptive ways (e.g. inflated public fear, unnecessary anxiety, and socio-economical unease).^7,10^ Governments should attempt to understand public emotions during a public health emergency and relay accurate information to ease the public’s fears.^8^ The power of social media is increasingly recognized in public health as a means of disseminating information.^11,12^ Social media surveillance can be used to assess the effectiveness of public risk communication, evaluate public reactions to existing interventions, and alert decision-makers about emerging rumours and misinformation.^13-16^ Furthermore, in contrast with traditional surveys, social media surveillance can systematically monitor public emotions and responses to outbreaks in real-time and is less likely to be affected by recall bias or reporting bias.^13,14^ Despite these benefits, social media surveillance during epidemic outbreaks has been under-explored, with only few studies using Twitter and Facebook having been conducted on A(H1N1) and Ebola.^14, 17-19^

With rapid advancement in mobile and search engine technology, e-commerce since 2010, China has its own social media platforms, which cover 95% of its residents and are a major part of their daily lives;^20^ however, there has been very little evidence of these platforms use in large-scale public health emergencies. Using social media surveillance data, this study aimed to assess real-time public attention, risk perception, emotion, and behavioural responses to live events during the first 11 weeks of the COVID-19 outbreak.

## Methods

### Data Collection

#### Time period

*1 Dec 2019-15 Feb 2020*

#### Baidu index

Baidu search engine, Google equivalent’s in China, has more than 1 billion Chinese users. The Baidu index, stemming from search frequency on the Baidu search engine, is powered by Baidu statistics and exhibited as pre-set keywords. The Baidu index reflects users’ attention on specific keywords over days, thereby assessing public awareness. For this study, we manually scanned and identified all Baidu index keywords that include COVID-19, recommended personal protection measures, and rumours and misinformation.

The National Health Commission issued a series of guidelines recommending residents adopt personal protection measures consisting of four main aspects: respiratory protection, hand hygiene, home disinfection and health monitoring. Accordingly, we identified “mask,” “hand sanitizer,” “disinfectant,” and “thermometer” as keywords to reflect the four above measures (Table 1). We also identified from the Weibo “Hot Search” ranking (see below Sina Weibo) three keywords: “radix isatidis,” “Shuanghuanglian,” and “garlic,” which were widely circulated rumours and misinformation regarding certain herbal medicines for personal health protection. For comparison, we also gathered Baidu data from last year during the same time-period as our baseline data: 1 Dec 2018-15 Feb 2019.

**Table 1:**
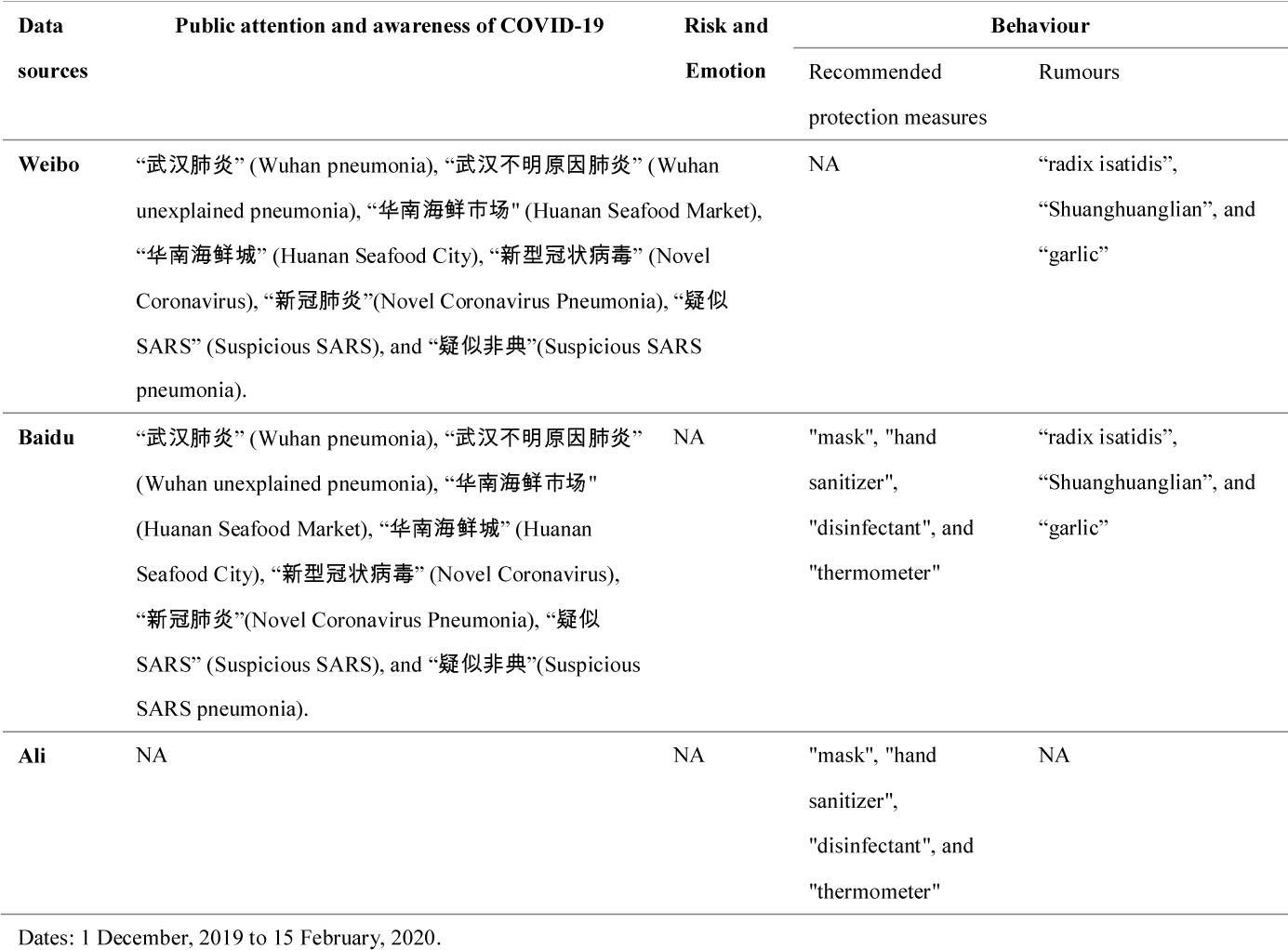
Keywords to measure public attention, risk perception, emotion, and behaviour

**Sina Weibo** (aka “microblog”) is a popular social networking site in China with more than 500 million registered users. Similar to Twitter, Sina Weibo is a free social media site that enables registered users to communicate with others in real-time using posts. We programmed a crawler in Python to collect Weibo statuses at the provincial level from the advanced search function of the Sina Weibo website from 1 Dec 2019 to 15 Feb 2020, collecting total of 6,216,572 Weibo posts (Appendix Figure 1). All posts gathered were sorted by date and user location (i.e. being either in or out of Hubei province). A list of Weibo keywords (hashtags##) about COVID-19 was manually scanned from the first five hundred identified posts and tested for sensitivity and specificity before being used for data crawling. Crawled Weibo posts were used, individually or in combination, to generate various Weibo indices of public attention and awareness on COVID-19, and rumours and misinformation (Table 1, Figures 1 and 5). Furthermore, in order to pinpoint events that triggered public reactions, we referred to Weibo’s “Hot Search” ranking to identify top Weibo topics that drew public attention (Appendix Table 1).

**Figure 1:**
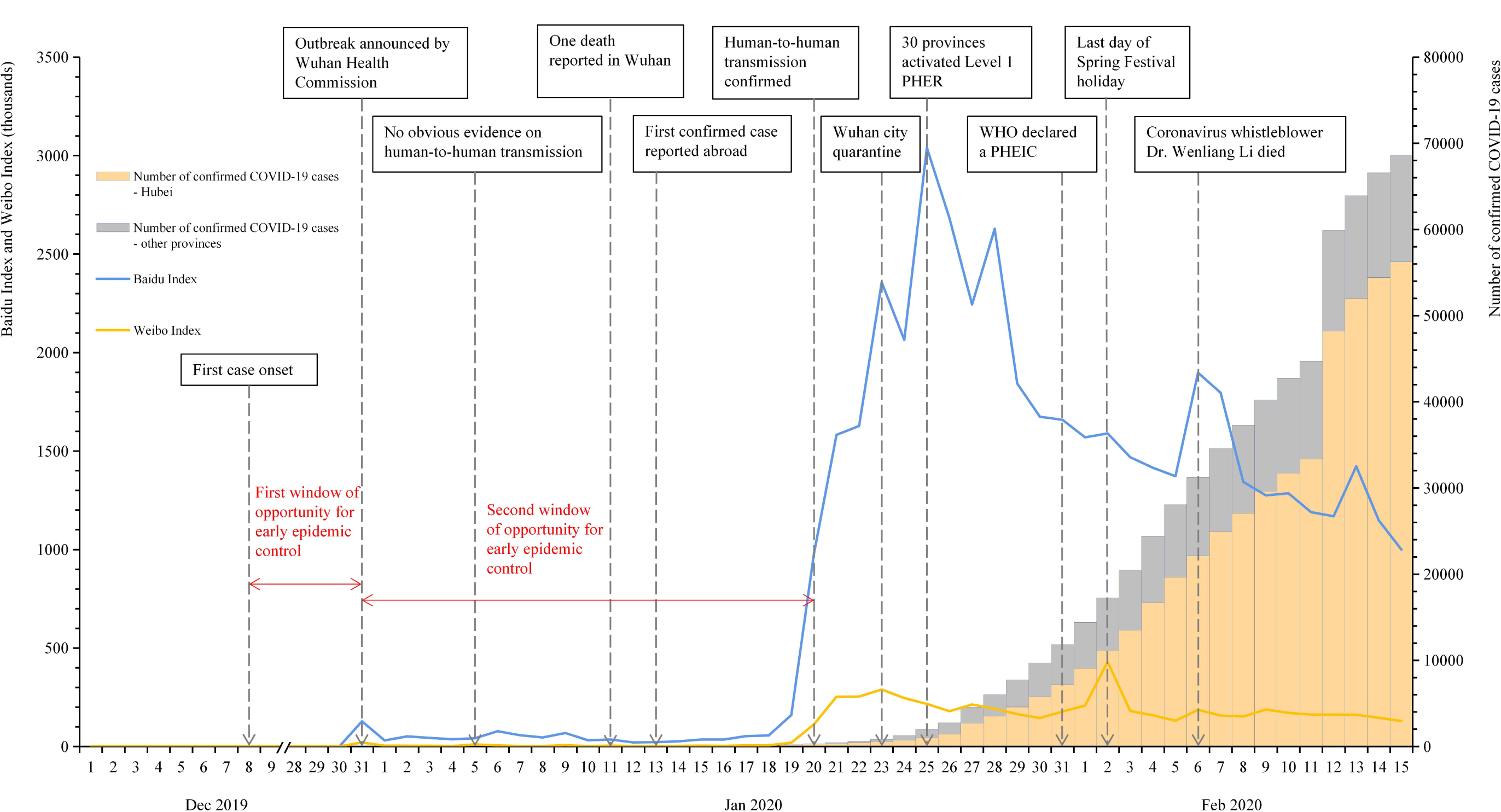
Public attention and awareness of COVID-19 by Baidu and Weibo index. Notes: Key events related to COVID-19 outbreak were identified on Weibo “Hot Search” ranking. PHER: public health emergency responses; PHEIC: public health emergency of international concern.

#### Ali index

Ali platform, Amazon’s equivalent in China, is a powerful and popular internet-based e-commerce marketplace with more than 1 billion users, and its purchasing index reflects the number of specific products purchased online. We employed the same keywords relevant to recommended personal protection measures: “mask,” “hand sanitizer,” “disinfectant,” and “thermometer,” to assess public behavioural responses to the COVID-19 outbreak (Understandably, Ali platform did not generate an index for rumour-related items). We similarly collected behavioural data during the current outbreak and in the year prior.

### Data analysis

***Public attention and awareness***, as well as **rumours and misinformation**, were assessed using both Weibo and Baidu daily indices. To assess public attention and awareness, we constructed a Weibo index using the daily number of “posts” with keywords related to COVID-19, where the proportions of Weibo posts that contained these keywords were calculated and presented on a daily basis. ***Public emotion*** was assessed by importing and processing all gathered Weibo posts in the Linguistic Inquiry and Word Count (LIWC) text analysis program, ^21,22^ for assessing psychological constructs, particularly relating to emotion. We adopted the Chinese version (SCLIWC), which has been previously adapted to the Chinese context and validated in Weibo posts.^23,24^ In this study, we assessed the elected psycholinguistic features: negative emotion (i.e. anxiety, sad, anger), and risk perception (i.e. drives). Examples of words included in these features are presented in Appendix Table 2. Using the Baidu and Ali daily indices, we assessed the ***intention and behaviours*** of adopting recommended personal protection measures and/or rumours about ineffective treatments during the COVID-19 outbreak.

Finally, we employed Spearman’s rank correlation analyses to detect the consistency of the Weibo, Baidu and Ali indices and to assess the correlation between different indices in different aspects of public response: public attention and awareness, risk perception and negative emotion, and intention and behaviours. We further conducted regression models to estimate the provincial differences in risk perception and various negative emotions across different periods.

## Results

### Public attention and awareness of COVID-19

The first COVID-19 case emerged on 8 Dec 2019, which was first announced by the Wuhan Municipal Health Commission on 31 Dec. The Baidu search index for COVID-19 had no results throughout the entire month of December, and jumped to 127,336 searches on 31 Dec 2019, when Wuhan Health Commission first reported a total of 27 patients with pneumonia of unknown cause (Weibo Hot Search #4). However, public attention and awareness immediately dropped to 30,504 searches the next day, and remained low between 1 and 19 Jan. Starting on 20 Jan, when the National Health Commission confirmed human-to-human transmission (Weibo Hot Search #1), it increased sharply and peaked at 3,039,324 on 25 Jan. It then steadily decreased with minor fluctuations. Figure 1 highlighted the squandered windows of opportunity for early epidemic control.

The Weibo index presented a similar trend as the Baidu index, with an exceptional peak on 2 Feb, the day before people were scheduled to return to work following the spring festival holiday (Weibo Hot Search #25). Correlation analysis demonstrated a positive correlation of public attention and awareness between the Baidu and Weibo indices (Spearman’s rank correlation coefficient 0.897, p < 0.001).

### Public risk perception and emotional response

A total of 2,937,082 Weibo posts were collected, including those in Hubei Province (159,624, 5.4%) and outside of Hubei (2,777,458, 94.6%). The daily proportions of Weibo posts expressing perceived risk and emotion (both positive and negative) are presented in Figure 2 and because public awareness of COVID-19 was low before 20 Jan, we observed strong fluctuations in the three indicators during this period caused by a small data size (many days Weibo posts <1000).

**Figure 2:**
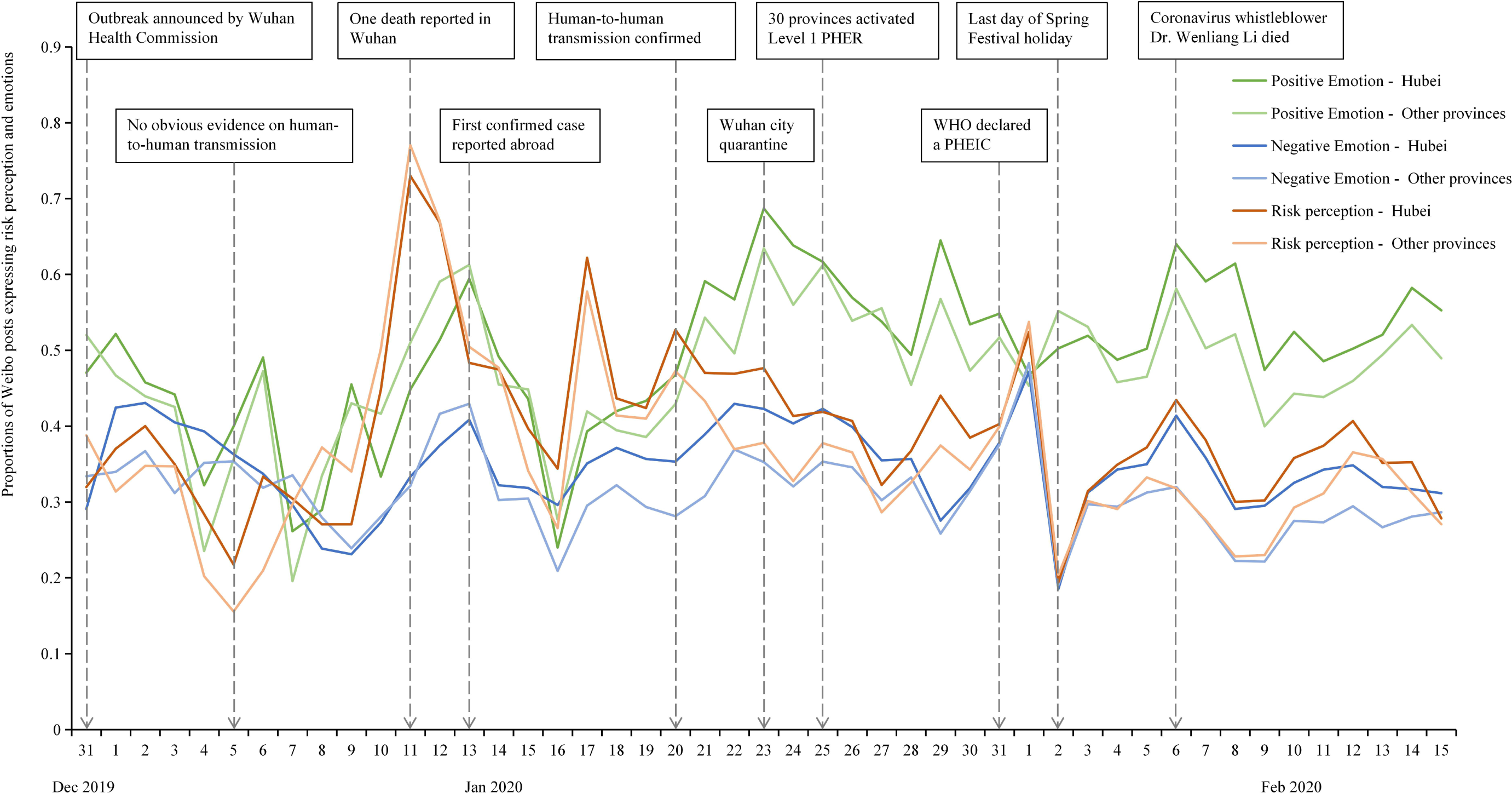
Daily proportions of Weibo posts expressing risk perception and emotions. Notes: PHER: public health emergency responses; PHEIC: public health emergency of international concern.

On 31 Dec 2019, 32.0% of Weibo posts (n=4,543) in Hubei province, where Wuhan is located, started to recognize the risk of this disease, and 29.1% expressed negative emotions. At the same time, 47% and 52% of Weibo posts in Hubei and other provinces, respectively, expressed positive emotions in celebration of the New Year 2020. Positive emotion has been consistently higher than negative emotion throughout the entire study period in Hubei and across China. On 3 and 5 Jan, the Wuhan Health Commission repeatedly announced “there was no obvious evidence of human-to-human transmission,” and confirmed that “the virus was neither severe acute respiratory syndrome coronavirus (SARS-CoV) nor Middle East respiratory syndrome coronavirus (MERS-CoV)” (Weibo Hot Search #4). Along with these announcements, the proportions of those expressing perceived risk and negative emotion showed steady reduction while positive emotion fluctuated across China for several days, signalling a period of confusion. On 11-13 Jan when one death from COVID-19 and the first confirmed case in Thailand were reported, respectively (Weibo Hot Search #29 and #7), the proportions of posts that expressed perceived risk and negative emotions reached a peak of around 75% and 40%, respectively, across all provinces in China. We observed a strong presence of public risk perception between the first death reported on 11 Jan and the confirmation of human-to-human transmission on 20 Jan. Since then, positive emotion took over and remained high in the 47-68% range in Hubei and 40-63% across China. One exception was that all three indicators merged on the day after 31 Jan, when WHO declared the outbreak as a PHEIC (Weibo Hot Search #7). On 2 Feb when public attention shifted to discussing whether work should resume the next day as originally planned, risk perception and negative emotion plunged sharply to a low-mark and remained low thereafter. Correlation analysis showed positive correlations between risk perception and negative emotion (Spearman’s rank correlation coefficient 0.500, p <0.001 for Hubei; 0.351, p=0.016 outside of Hubei).

Figure 3 presents a more detailed analysis of the three negative emotions expressed on Weibo: public anxiety, sadness and anger; all reached their peak three times around 1 Jan, 13 Jan, and 6 Feb, and manifested within 24 hours after the triggering event took place. Reports of the first confirmed case abroad triggered more anxiety, sadness and anger outside of Hubei province, and risk perception about imported cases from Hubei rose. On 13 Jan, 18.0% of Weibo posts outside of Hubei were angry with the outbreak, much higher than 8.3% in Hubei. On 6 Feb, Wenliang Li, the whistle-blowing doctor who first warned of the outbreak and who was accused by police of spreading fake news, died from COVID-19.^25^ This renewed public expression of negative emotions, especially in Hubei. Around 23% of Weibo posts in Hubei expressed anxiety, sadness and anger in response to Dr. Li’s death, much higher than the 14% of users outside of Hubei.

**Figure 3:**
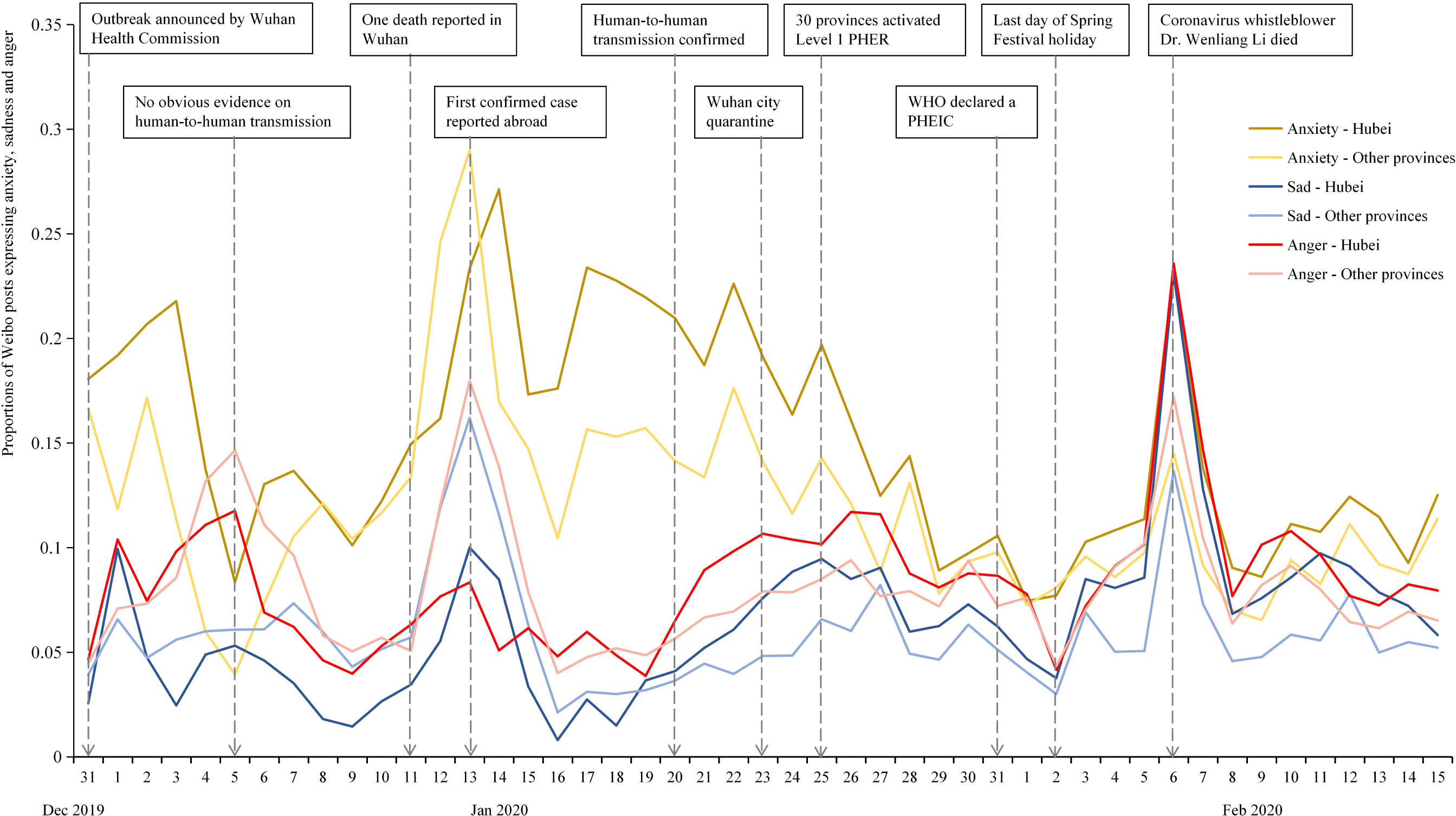
Daily proportions of Weibo posts expressing anxiety, sadness and anger. Notes: PHER: public health emergency responses; PHEIC: public health emergency of international concern.

It is worth noting that the confirmation of human-to-human transmission on 20 Jan marked the beginning of a steady decline in public anxiety; during the same period, public sadness and anger increased, but started to decline after the implementation of containment measures on 23-25 Jan. Wuhan city implemented a series of epidemic control and quarantine measures starting 23 Jan: public transportation in the city was shuttered, and all flights and trains leaving from Wuhan were cancelled (Weibo Hot Search #13). On 25 Jan, 30 provinces (autonomous regions, municipalities) in mainland China, excluding Tibet, activated major public health emergency responses (Weibo Hot Search #11). To assess the impact of the 20 Jan confirmation of human-to-human transmission on public perceived risk and negative emotions, we conducted regressions for the subsamples before and after the communication (Table 2), and found that there was no evidence suggesting differences in public perceived risk and all three negative emotions between residents in Hubei and outside of Hubei before 20 Jan; however, evidence showed Hubei residents were significantly more likely to have higher perceived risk and negative emotions than those outside of Hubei after 20 Jan.

**Table 2:**
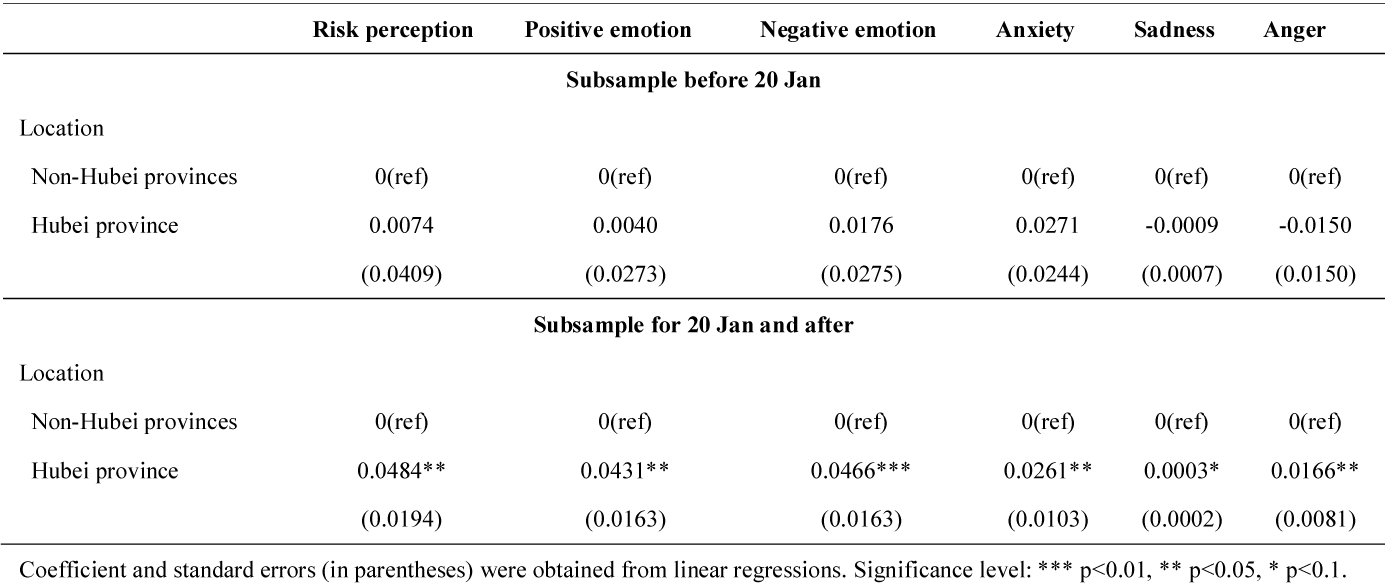
Association between location and emotional responses separately before and after 20 Jan

### Behavioural Responses

National Health Commission first proposed respiratory protection and hand hygiene in the protection guidelines issued on 21 Jan, and first proposed the use of home disinfection and health monitoring on 22 and 25 Jan, respectively. The public responded quickly to the issuing of specific guidelines, and both intended and actual purchasing behavior dramatically increased accordingly. The Baidu and Ali indices of the four recommended personal protection measures, except for Ali index of thermometer, all started to increase sharply on 21 Jan, with an exceptional drop on 24 Jan (Chinese New Year’s Eve) (Figure 4). The Ali index of thermometer increased rapidly from 25 Jan. The Baidu index indicating an intention to adopt all four measures increased earlier than the actual behaviors presented in the Ali index. Both indices were steadily low during the same period last year (baseline).

**Figure 4.**
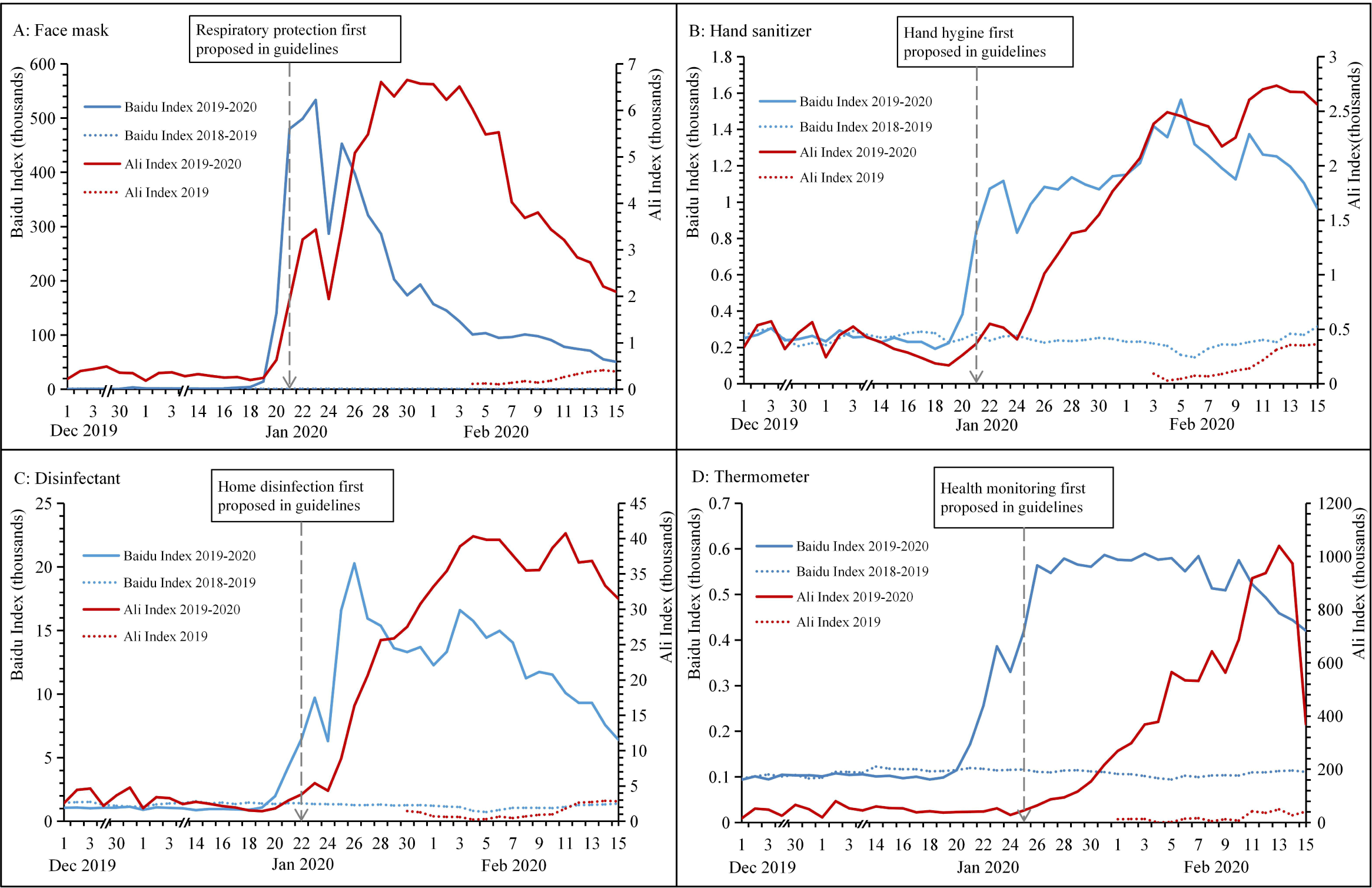
Trend of Baidu and Ali index on recommended personal protection measures. Notes: Ali index can be only available within one year, with no data on Dec 2018 and Jan 2019.

Figure 5 shows trends in the Baidu and Weibo indices on rumors, representing intended behavior and public attention, respectively. Radix Isatidis is a traditional Chinese medicine for fever, and its effectiveness is still widely believed in China. The Baidu index of “radix isatidis” started to increase when the outbreak was first announced. It further increased sharply on 20 Jan, when human-to-human transmission was confirmed, and reached a peak on 21 Jan. It started to decline from 21 to 24 Jan, during which the People’s Daily issued three reports in an attempt to refute rumors of its effectiveness against COVID-19. On 31 Jan, the People’s Daily reported that Shuanghuanglian, another Chinese medicine, could inhibit COVID-19, and both Baidu indices of “Shuanghuanglian” and “radix isatidis” rapidly reached their peaks. Both indices decreased rapidly on 2 Feb, when the People’s Daily clarified this misinformation, stating Shuanghuanglian cannot prevent COVID-19. The rumor that “garlic can prevent COVID-19” started to spread on Weibo on 21 Jan, and the Baidu index of “garlic” increased accordingly. It reached a peak on 27 Jan, and declined after 28 Jan, when the People’s Daily first refuted rumors about the protective function of garlic. However, these three Baidu indices were still higher than their previously observed levels, even during late Feb. Representing the baseline, these indices had been steadily low during the same period last year. The Weibo index, representing public attention, demonstrated the same pattern as the Baidu index, representing intended behavior, for rumors.

**Figure 5.**
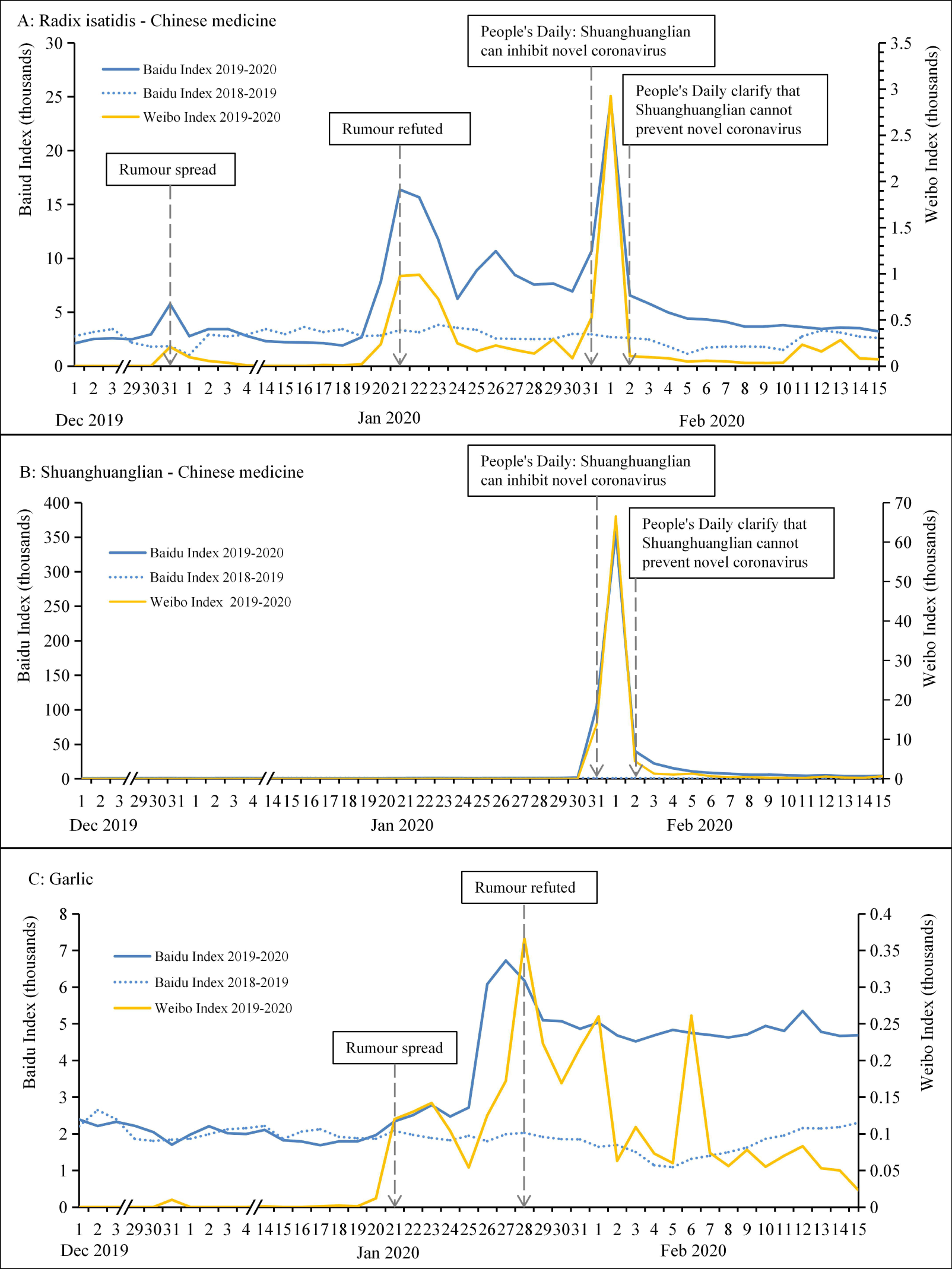
Trend of Baidu and Weibo index on rumors.

We further conducted correlation analysis (Appendix Table 3), which showed strong positive correlations between the Baidu and Ali indices for all recommended measures (correlation coefficient range of 0.676-0.860, p <0.001) and positive correlations between the Baidu and Weibo indices for all rumors (correlation coefficient range of 0.765-0.916, p <0.001).

## Discussion

This study is the first social media surveillance analysis to track and assess real-time public emotional and behavioural responses to government announcements, rumours and misinformation, and major epidemiological events during the first 11 weeks of the COVID-19 outbreak in China. Delayed release of information ignited negative public sentiment. Public risk perception and anxiety started to decline after the confirmation of human-to-human transmission, whereas sadness and anger increased until the implementation of containment measures. We found the public responded quickly to government announcements and followed official guidelines accordingly, but rumours and misinformation regarding herbal remedies were found to have led to irrational panic buying during the outbreak. The timely clarification of rumours effectively reduced irrational behaviour.

Compared with traditional surveys, social media surveillance provides data that are real-time, longitudinal, and dynamic in capturing public attention/awareness, risk perception, emotions, intentions, and behavioural reactions, and can be an effective means to assess government’s risk communication efforts and assist in epidemic control and response. This study presented a rigorous methodological process for conducting social media surveillance during epidemic outbreaks that can inform future studies, especially in the Chinese context. There are several limitations. First, there is censorship on Weibo in China, and thusly there was a possibility that we might not have captured all posts. To reduce this bias, we collected data (i.e. Weibo posts) daily, before government intervention. Second, LIWC is a tool that detects emotions using the frequency of words instead of the whole posts, which may lead to misclassification. We adopted its adapted Chinese version, which had been validated locally and can better match with Weibo to reduce misclassification. Third, there is an inherent bias in social media studies where data might misrepresent the real world because users might present themselves differently online and/or represent a skewed population towards the young.

The lack of transparent, timely, and effective risk communication from health authorities on an emerging infectious disease in its early stages failed to bring about the appropriate level of public awareness and behavioral responses, such as avoidance of mass gatherings. The government did not provide any actionable advice for personal protection until 21 Jan. There were two missed windows of opportunity for early epidemic control of COVID-19 that we identified during the first 11 weeks of the outbreak: 1) the first COVID-19 case emerged on 8 Dec 2019, more than three weeks before 31 Dec, when the Wuhan Health Commission finally announced the outbreak;^1-3^ 2) between 31 Dec and 19 Jan, the Wuhan Health Commission made four public announcements emphasizing that “there was no obvious evidence for human-to-human transmission” without providing any actionable advice on personal health protection, and the eight physicians who attempted to raise awareness of the outbreak had been warned by the police to keep silent.^25^ This series of government announcements kept public attention and awareness low, preventing the public from realizing the risk of the disease and from taking personal protection measures at an earlier time. This period overlapped with the celebration of the arrival of 2020 and preparation for the Chinese New Year (25 Jan), the most important period of the year for family reunion and social gathering in China. Instead of raising public awareness about the possible outbreak, on 18 Jan local Wuhan officials and residents held a traditional social gathering, where tens of thousands of residents attended a joint celebration banquet that was promoted on the front-page of local newspapers.^26^

Despite uncertainty and incomplete information about the risk, the early release of information on emerging infectious diseases and early implementation of containment measures might effectively address public sentiment and control the COVID-19 epidemic and future outbreaks. Our data showed public risk perception and negative emotions were highly correlated to outbreak news events, government announcements, and the implementation of containment measures. Acknowledging the knowns and unknowns about an emerging health threat and taking proactive mitigation measures help ease public fear and build public trust. Our previous studies showed that, although the prevalence of anxiety was high at 33% among Wuhan residents, 92% supported quarantining Wuhan, and 75% were confident that governmental containment measures would quell the outbreak.^28-29^ Interestingly, the proportion of Weibo posts that expressed positive emotion were significantly higher than negative emotion throughout the entire study period, especially after the implementation of restrictive public health measures. Looking closer at the data showed the positive emotion was boosted by a strong sense of solidarity for Wuhan and for health care workers who worked at the frontline. The timing of the lockdown coincided with the Chinese New Year with unexpected extended family time also lifting public spirits.

By incorporating social media in the emergency response system, health authorities can detect misinformation, confront inaccurate messages in real-time and assess the effectiveness of current epidemic control measures for immediate improvement. ^30^ As an example, during this outbreak, several incidents of panic buying were triggered by both official guidelines and rumours (e.g. masks, Shuanghuanglian, or garlic), and these behaviours were dramatically reduced after clarification. The most popular social media platforms in China, such as WeChat and Weibo, and several health professionals’ online communication platforms (e.g. Dingxiangyuan) established their own platforms to clarify rumours. Working with the private sector to ensure sufficient stock and reasonable pricing for recommended personal protection products, such as hand sanitizer and masks, is critical for strengthening epidemic prevention and control and avoiding public panic. As we collected social media data, we found that the public shared information about their disease symptoms, their health care seeking experiences, and the social/cultural aspects of risks and containment strategies. Social media surveillance has the potential to track both disease activity before it reaches health care facilities and public concerns and risk perceptions as they emerge and evolve in real time.^13-15^ Future research should explore the epidemiological information shared on these platforms and examine the role of social media in early detection and early warning during epidemic preparedness and response.

## Conclusion

During an outbreak, real-time social media surveillance can enable a timely assessment of public reactions to risk communication and epidemic control measures and the timely clarification of rumors; as such, it should be fully incorporated into epidemic preparedness and response systems.

## Data Availability

All data are publicly available.

## Contributors

Z.H. and L.L. conceptualized the study design. X.Z., H.J., F.D. collected and analysed data. Z.H., L.L., F.D, H.J. interpreted results and wrote the manuscript. All authors approved the final draft of the manuscript.

## Ethics

All data are publicly available.

## Acknowledgements

Z.H. acknowledges financial support from the National Natural Science Foundation of China (No. 71874034).

---

**Appendix Figure 1.**
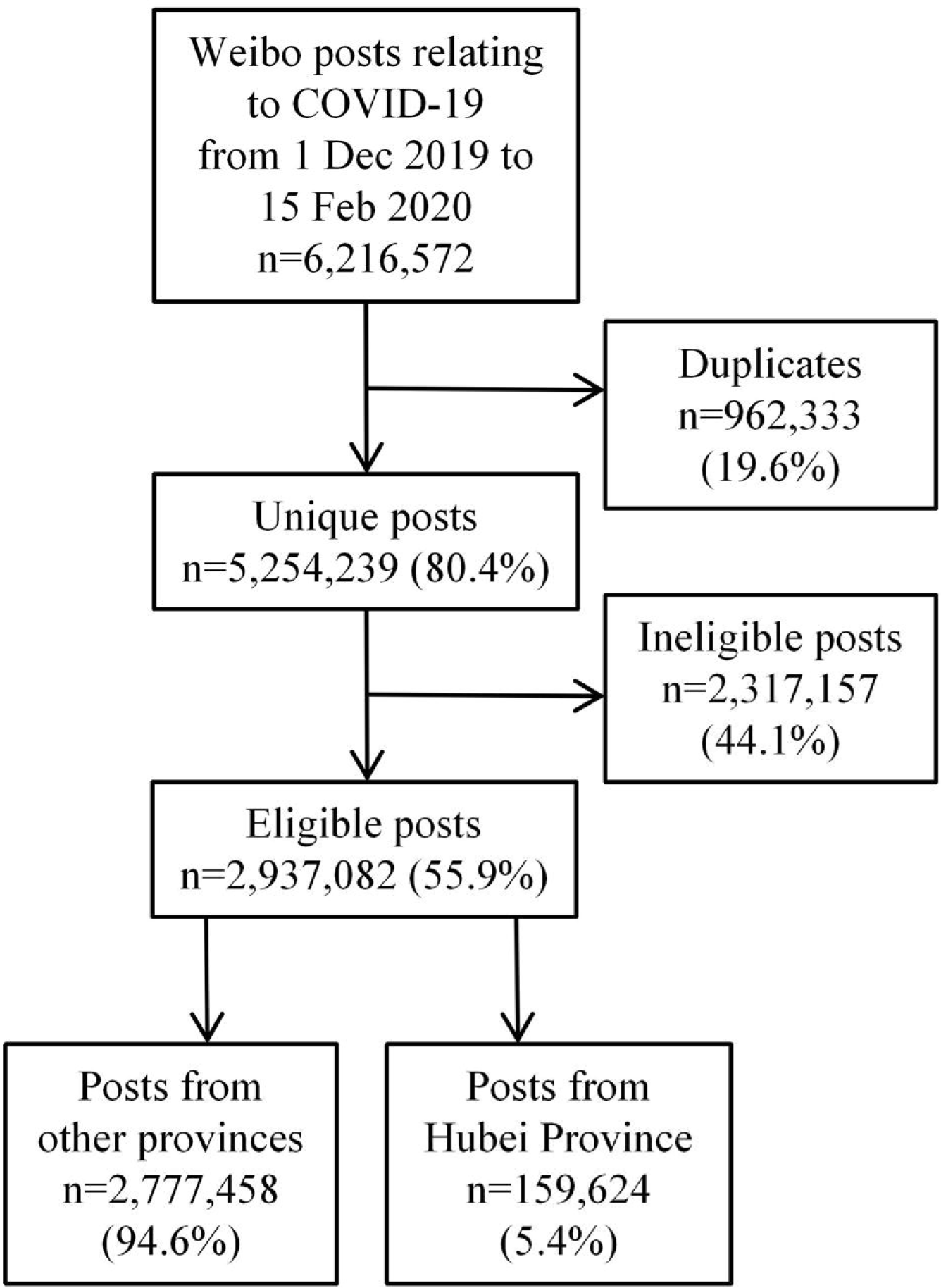
Flowchart for Weibo posts collecting process. Notes: Ineligible posts: A Python program is designed to exclude status messages that were not considered to be public opinion expression such as news, advertisements, and government announcements, etc.

## References

1. Huang C, Wang Y, Li X, et al. Clinical features of patients infected with 2019 novel coronavirus in Wuhan, China. Lancet 2020; 395(10223): 497–506.

2. Chen N, Zhou M, Dong X, et al. Epidemiological and clinical characteristics of 99 cases of 2019 novel coronavirus pneumonia in Wuhan, China: a descriptive study. Lancet 2020; 395(10223): 507–513.

3. Li Q, Guan X, Wu P, et al. Early Transmission Dynamics in Wuhan, China, of Novel Coronavirus-Infected Pneumonia. N Engl J Med 2020.

4. National Health Commission of the People’s Republic of China. http://www.nhc.gov.cn. (accessed Mar 2, 2020).

5. Wu Z, McGoogan JM. Characteristics of and important lessons from the Coronavirus Disease 2019 (COVID-19) outbreak in China: summary of a report of 72□314 cases from the Chinese Center for Disease Control and Prevention. JAMA 2020.

6. Lin L, Savoia E, Agboola F, Viswanath K. What have we learned about communication inequalities during the H1N1 pandemic: a systematic review of the literature. BMC public health 2014; 14: 484–8.

7. Lin L, Jung M, McCloud RF, Viswanath K. Media use and communication inequalities in a public health emergency: a case study of 2009-2010 pandemic influenza A virus subtype H1N1. Public health reports (Washington, DC : 1974) 2014; 129 Suppl 4(Suppl 4): 49–60.

8. The Lancet. COVID-19: fighting panic with information. Lancet 2020; 395(10224): 537.

9. Brooks SK, Webster RK, Smith LE, et al. The psychological impact of quarantine and how to reduce it: rapid review of the evidence. Lancet 2020. pii: S0140-6736(20)30460-8.

10. John Sumo et al. Risk and Outbreak Communication: Lessons from Taiwan’s Experiences in the Post-SARS Era. Health Secur 2017; 15(2): 165–169.

11. St Louis C, Zorlu G. Can Twitter predict disease outbreaks? BMJ 2012; 344: e2353.

12. The Lancet. The medium and the message of Ebola. Lancet 2014; 384: 1641.

13. Gabriel J Milinovich, Gail M Williams, Archie C A Clements, Wenbiao Hu. Internet-based surveillance systems for monitoring emerging infectious diseases. Lancet Infect Dis 2014; 14(2): 160–8.

14. Tang L, Bie B, Park SE, Zhi D. Social media and outbreaks of emerging infectious diseases: A systematic review of literature. Am J Infect Control 2018; 46(9): 962–972.

15. Mollema L, Harmsen IA, Broekhuizen E, et al. Disease Detection or Public Opinion Reflection? Content Analysis of Tweets, Other Social Media, and Online Newspapers During the Measles Outbreak in the Netherlands in 2013. J MED INTERNET RES 2015; 17(5): e128.

16. Oyeyemi SO, Gabarron E, Wynn R. Ebola, Twitter, and misinformation: a dangerous combination? BMJ 2014; 349: g6178.

17. Fung IC, Duke CH, Finch KC, Snook KR, Tseng PL, Hernandez AC, Gambhir M, Fu KW, Tse ZTH. Ebola virus disease and social media: A systematic review. Am J Infect Control 2016; 44(12): 1660–1671.

18. Fung IC, Tse ZT, Cheung CN, Miu AS, Fu KW. Ebola and the social media. Lancet 2014; 384(9961): 2207.

19. Signorini A, Segre AM, Polgreen PM (2011). The Use of Twitter to Track Levels of Disease Activity and Public Concern in the U.S. during the Influenza A H1N1 Pandemic. PLOS ONE 2011; 6(5): e19467.

20. Isaac Chun-Hai Fung et al. Chinese Social Media Reaction to Information about 42 Notifiable Infectious Diseases. PLOS ONE 2015; 10(5): e126092.

21. Pennebaker J, Boyd R, Jordan K, Blackburn K. The development and psychometric properties of LIWC2015, Austin, TX: University of Texas at Austin; 2015.

22. Tausczik Y, Pennebaker J. The psychological meaning of words: LIWC and computerized text analysis methods. J Lang Soc Psychol 2010; 29:24–54.

23. 黃金蘭, Chung C K, Hui N, et al. 中文「語文探索與字詞計算」詞典之建立 中華心理學刊 2012, 54(2):185–201.

24. Zhao N, Jiao D, Bai S, et al. Evaluating the Validity of Simplified Chinese Version of LIWC in Detecting Psychological Expressions in Short Texts on Social Network Services. PLOS ONE 2016, 11(6):1–15.

25. West China Metropolis Daily. From “rumor maker” to “respectable person” Doctor Li Wenliang’s 2020 New Year. http://news.jxntv.cn/2020/0203/9329454.shtml/ (accessed Feb 3, 2020).

26. Chutian Metropolis Daily. More than 40,000 families gathered for a group dinner to celebrate Chinese New Year at Baibuting community in Wuhan. https://ctdsbepaper.hubeidaily.net/pc/column/202001/19/node_A03.html/ (accessed Jan 19, 2020).

27. Gu H, Chen B, Zhu H, Jiang T, Wang X, Chen L, Jiang Z, Zheng D, Jiang J. Importance of Internet surveillance in public health emergency control and prevention: evidence from a digital epidemiologic study during avian influenza A H7N9 outbreaks. J Med Internet Res 2014; 16(1):e20.

28. Hou Z, Lin L, Lu L, et al. Public Exposure to Live Animals, Behavioural Change, and Support in Containment Measures in response to COVID-19 Outbreak: a population-based survey in China. medRxiv 2020.02.21.20026146.

29. The Lancet Infectious Diseases. Challenges of coronavirus disease 2019. Lancet Infect Dis 2020;20(3):261.

30. Depoux A, Martin S, Karafillakis E, Bsd RP, Wilder-Smith A, Larson H. The pandemic of social media panic travels faster than the COVID-19 outbreak. J Travel Med. 2020. pii: taaa031. doi: 10.1093/jtm/taaa031.

